# Effect of a 12-mo milk-based micronutrient-fortified drink intervention on children: a systemic analysis of placebo-controlled study dataset

**DOI:** 10.1101/2022.05.03.22273437

**Authors:** Karan Lomore, Vani Gangwar, Samreen Rizvi, KV Venkatesh

## Abstract

**Background:** Nutritional deficiencies have many immediate and long-term effects on physical and cognitive development outcomes, with the children not achieving their full potential. A milk-based health drink fortified with micronutrients as a part of a daily balanced diet can promote physical and cognitive growth in children by increasing macronutrients and micronutrients availability in the body. The systematic analysis aims to quantify the effect of a formulated health drink on children’s physical, clinical, and cognitive development outcomes.

**Methods:** The dataset used in the analysis was obtained from literature and consisted of 900 children between 7 to 12 years of age. These children were distributed equally into the Control group (no micronutrient-fortified health drink is given), Group I (micronutrient-fortified health drink in water), and Group II (micronutrient-fortified health drink in the toned milk).

**Results:** The analysis shows that micronutrient-fortified health drinks in water (by 2.1-fold) and toned milk (by 2.5-fold) improve the height gain velocities and anthropometric and body composition parameters. It helped children achieve healthy IAP growth percentiles compared to the control group. The analysis also shows a 1.6- and 2-fold change in the cognitive tests grades in groups I & II, respectively, compared to the control group.

**Conclusions:** The analysis indicates that two servings of 33g micronutrient-fortified health drink in water and toned milk for a year significantly improved the gain velocities of anthropometric and body composition parameters, reduced time taken to complete the physical activity task, reduced the number of anaemia and morbidity cases among groups, and improved the scoring grades in cognitive assessment test in studied children population. The IAP data benchmarking clearly indicated that a micronutrient fortified milk-protein-based powder significantly improved children’s overall growth with better health.

## Background

The importance of proper growth and development defines a need for a balanced diet that ensure adequate daily intake of nutrients such as macro and micro-nutrients as per the recommended dietary intake for a particular age group. Macronutrients are the primary energy source for the human body during the growth and maintenance phase. These are mainly categorised into carbohydrates, proteins, and fats. Micronutrients include vitamins and minerals important for optimal growth and maintenance of the body functions [1]. Good daily nutrition supports optimal physical and cognitive growth and development, influencing an individual to perform tasks efficiently [2]. The optimal macro and micro-nutrient rich diet also boost immunity, consequently lowering the chances of common morbidities and improving clinical health [3]. An inadequate dietary intake can hinder proper growth during development [4]. A child’s growth and development are very rapid in a specific time. A rapid increase in the velocity of height and weight with cognitive development is observed in children during their second growth spurt. Therefore, substantial emphasis should be given to balanced nutrition during these times to achieve optimum growth [5]. A recent report by UNICEF indicates that 50% of the surveyed adolescents (10 -19 yrs.) in India suffer from at least two of the six micronutrient deficiencies, including Iron, Folate, Vitamin B12, D, A and zinc [6].

Several interventions and supplementation studies have evaluated the importance and benefits of multiple nutrients on various growth parameters, including physical growth, serum micronutrient status, cognitive functions, and clinical outcomes in the Indian population. Studies have shown the effect of micronutrient supplements on physical performance outcomes, including endurance, speed, aerobic capacity, and visual reaction time in children of both genders [2]. It has been studied that micronutrient supplementation with high-quality milk protein intervention improves height in school-going children (6 -16 years old) and increases linear growth [7-10]. The effect of multiple micronutrient supplementation on children’s growth and nutritional status is also studied in terms of improved serum micronutrient status of Iron, Vitamin C, B12, A, and D [11, 12]. Multi-micronutrient status in young Indian children has been studied to investigate the role of micronutrients on haemoglobin concentrations and anaemia and provide evidence of lowered morbidity from diarrhoea and respiratory infections and has positive effects on growth and cognitive functions [13].

Several studies have shown that linear growth and short-term memory improvement can be achieved by administering higher micronutrient treatment with Iron, Iodine, Zinc, Omega-3 PUFAs, Vitamin A, B6, B12, Folate, Riboflavin, and Iodine in Indian children aged between 6-10 years. These nutrients are important for proper brain development and cognitive functions [14]. The micronutrients also affect other growth parameters, including changes in body weight, MUAC, and cognitive functions, such as retrieval ability and cognitive speediness in school children aged 6-15 years [15]. A 14 months placebo-controlled study showed a significant effect of a daily micronutrient-rich intervention on children’s cognitive functions such as attention and concentration [16]. Meta-analysis on the effect of multiple micronutrient interventions in healthy children indicated an association between micronutrients, academic performance, along with an increase in fluid intelligence leading to intellectual development [17]. Iron and Selenium influence has been studied to design effective intervention programmes for controlling micronutrient status and addressing growth, cognitive development and lowered morbidity in children [18, 19]. Therefore, a correct balance of nutrients becomes essential for a child’s optimal development.

After a detailed literature review, it becomes evident that there is an opportunity to introduce a comprehensive approach to gather holistic insights capturing the effect of nutrients on overall growth and development. Most studies focus on either one or two major areas for analysis, including physical performance, anthropometric parameters, morbidity, clinical outcomes, or cognitive development, but none gathered a comprehensive study on these major responses during a child’s growth years. It also becomes clear that multi-nutrient interventions are considered effective, and most studies are based on the statistical summary. Therefore, an intervention or supplement is an effective solution for preventing and addressing nutrient deficiencies in specific at-risk groups, as its regular dose or serving can satisfy daily nutrient requirements [20].

The present analysis utilises the dataset from the previous 12-month placebo-controlled health drink intervention study in 2008 to combat childhood nutritional status and its effect on overall growth and development. The study focused on evaluating the efficacy of a milk-based health drink in improving the different physical and cognitive growth parameters through statistical analysis and clinical outcomes using blood haemoglobin levels [21, 22]. The analysis provided critical insights for only a specific timespan which becomes difficult to extrapolate in further studies. Also, updated population-specific reference growth charts are needed to assess a child’s proper growth in recent times. The Indian Academy of Pediatrics (IAP) produced and recommended IAP 2015 Growth charts for monitoring Indian children. In 2006, the World Health Organization (WHO) published the first global growth standards for children under the age of 5 years, and the Government of India and IAP have adopted these standards for use in under five Indian children. Growth patterns differ among different world populations in older children due to nutritional, environmental, and genetic factors. Thus, IAP growth charts were developed using recent studies on children’s growth, nutritional assessment, and anthropometric data of healthy Indian children between the age of 5 and 18 from a total of 87022 (M: 54086, F: 32936) of upper and middle socioeconomic classes, where height, weight, and age were available for every child [23].

Therefore, the current study focuses on analysing the effect of health drink on the change in the gain velocity of a given anthropometric and growth parameters such as physical growth, physical performance time, clinical outcomes, and cognitive functions along with the body composition parameters such as body fat mass, fat-free mass, and bone mineral content. The strength of the present analysis is to benchmark growth parameters on new IAP growth charts to establish the validity of data in today’s context. Using probability distribution and new visualisation techniques, these longitudinals rather than cross-sectional growth references better represent various growth parameters changes upon multi-nutrient intervention.

## Methods

### Dataset description

Information of 900 school children from middle- and low-income families were obtained from available literature [21, 22]. The ethical approval for the original studies was obtained from the Avinashilingam University’s Ethical Committee (HEC.2006.01). The parents of all the participating children were informed about the study and written consent was obtained. The data consisted of children (i) that lied in the range of 7-12 years, (ii) boys to girls’ sex ratio was 1.04, and (iii) all children in the dataset were clinically healthy and were not at the risk of severe under-nutrition and severe obesity as per the IAP growth standards [23]. The data consisted of three groups, including one control group and two experimental groups, where one group was administered health drink dissolved in water and the other in toned milk, containing 300 children each with 50 children lying in the age group of 7+ to 12+ years. The Control group was not given any health drink. 54 dropouts were not included in the analysis. For each child, information relating to (i) baseline, mid, and final weight measurements, (ii) height, (iii) mid-upper arm circumference (MUAC), (iv) haemoglobin, (v) anaemia, and (vi) morbidity values were provided. The time taken by a child to complete a physical activity task was also documented. The scores and grades of cognitive tests such as Raven’s coloured progressive matrices (RCPM) test, Digit Span, Arithmetic Test, and Digit Symbol were also available for analysis. Refer Additional file for detailed methodology.

### Intervention

For this systemic analysis study, we considered the following as intervention and control groups:

1. Group I received 33 grams of health drink mixed in 150 ml of water twice a day and their usual daily diet.
2. Group II received 33 grams of health drink mixed in 150 ml of toned milk twice a day in addition to their usual daily diet.
3. The Control group received their usual daily diet without any health drink.

The daily diet was estimated with a 24-hour diet recall survey in 10 per cent of total children (n=90) through stratified random sampling [22]. The nutrient composition of the micronutrient-fortified formulation with water and toned milk is provided in the S1 Table.

### Assessment

The analysis represents the effect of 12 months of micronutrient-fortified formulation on various parameters of physical growth, clinical outcomes, and cognitive function in the Control, Group I and II. The anthropometric and body composition parameters were analysed with a change in gain velocity (rate) due to intervention. The change in the percentage of children under a certain IAP percentile was also calculated at baseline and at the end of the study. The data associated with clinical outcomes and cognitive function tests were also analysed. Descriptive analysis and visualisation techniques were used to compare and visualise the dataset of all three groups. The Mann–Whitney U test was used to determine the statistical significance (p-value<0.05) of intervention among Control and experimental groups. A detailed explanation of the further analysis is provided in Additional file.

### Physical growth analysis

The physical growth analysis includes anthropometric parameters like weight and height, MUAC, BMI and physical performance time. The body composition parameters included are fat mass, fat-free mass, and bone mineral content (BMC). The fat mass, fat-free mass, and BMC were not assessed in the original study dataset [21]. These parameters were derived using correlations mentioned in Additional file 1.

### Clinical analysis

The clinical analysis includes measurements related to anaemia and morbidity within the participants.

### Cognitive development analysis

The cognitive development analysis consisted of various tests such as Raven’s Coloured Progressive Matrices (RCPM) and Malin’s Intelligence test, which included Arithmetic test, Digit Span, and Digital Symbol tests for children. These tests were used to test the improvement in intelligence, numerical ability, memory, and concentration, respectively.

## Results

### Physical growth analysis

The anthropometric measurements determined the height and weight gain velocities (Fig 1). Fig 1A shows a 1.82- and 2.53-fold higher weight gain velocity in Group I and II, respectively, compared to the Control group (0.17 ± 0.07 kg/month). This resulted in 27% and 48% of children moving to above 50^th^ percentile in Group I and Group II, respectively, compared to only 12% of children moving above 50^th^ percentile in the Control group, as characterised by IAP data. The probability distribution (as shown in S1A Fig) demonstrates that 83% of children had greater weight gain velocity in Group I and 98% in Group II with higher gain velocity than the mean value seen in the Control (i.e., 0.17 kg/month). Further, no gender bias was observed since the distribution overlapped for boys and girls (S1B-S1C Figs). The Mann–Whitney U test for weight velocities of Group I & II compared to Control have shown statistical significance (p-value<0.05) for all observed outcomes.

**Fig 1:**
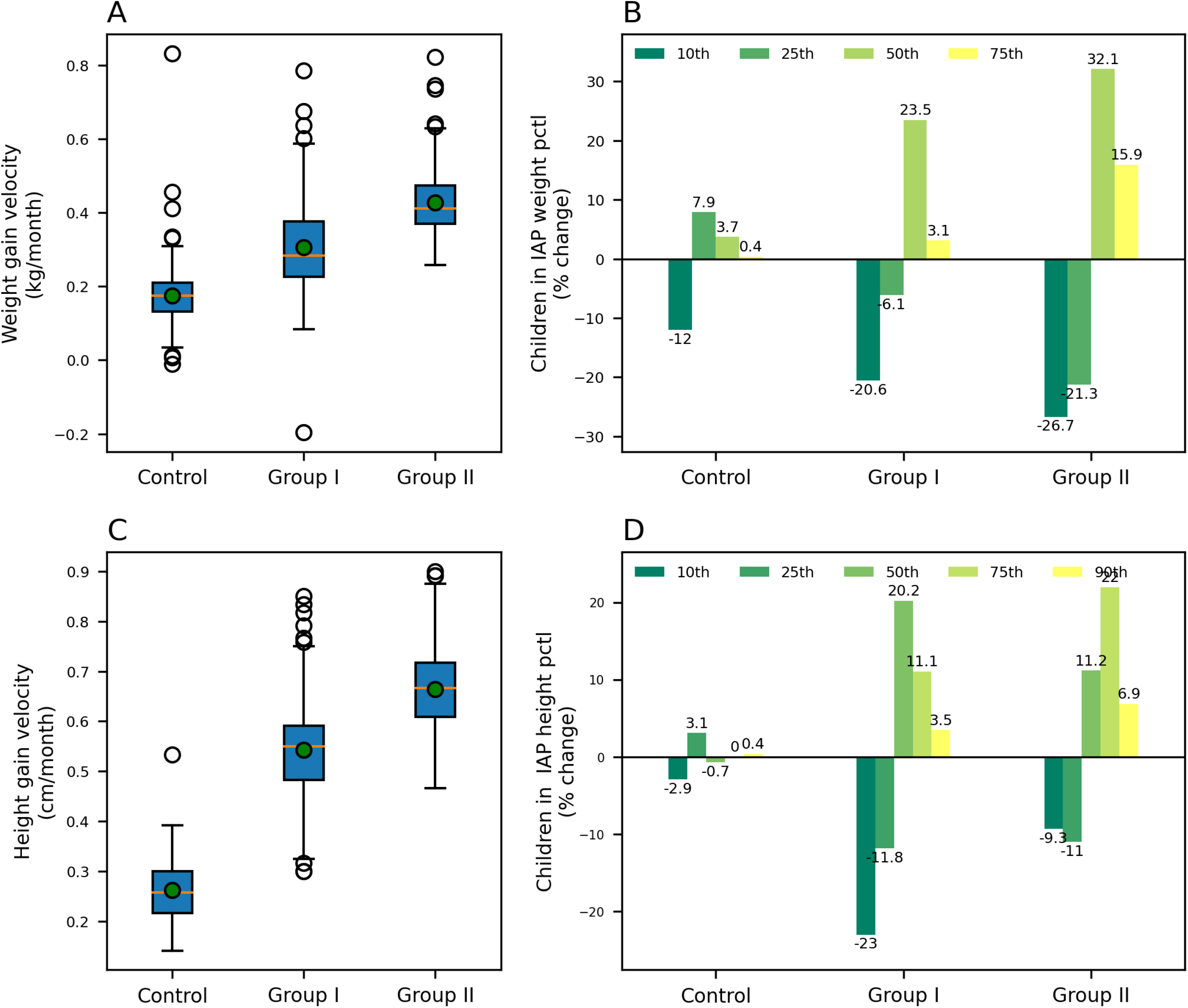
Weight and Height gain velocities and percentage change in IAP percentile children in Control, Group I and II. (A) Weight gain velocity, (B) Percentage change in IAP weight percentiles and (C) Height gain velocity and (D) Percentage change in IAP height percentiles for Control, Group I & II. The weight gain velocities for Group I, II and Control were (0.31 ± 0.12), (0.43 ± 0.08) and (0.17 ± 0.07) kg/month respectively, while the height gain velocities for Group I, II and Control were (0.54 ± 0.10), (0.66 ± 0.08) and (0.26 ± 0.06) cm/month, respectively. The green dot indicates that A and C represent the mean values. The darker green to yellow colour bars in B and D exhibit the increasing order of percentiles.

Fig 1C shows the height gain velocities, wherein 2.08- and 2.54-fold higher velocities were seen for Group I and Group II, respectively, relative to the Control (0.26 ± 0.06 cm/month). This resulted in 35% and 20% of the children moving to above the 50^th^ percentile in Group I and Group II, respectively, compared to only 3% in the case of the Control group, as characterised by IAP data. Although Group I demonstrated an overall higher velocity in height gain, the percentage of children moving to above 75^th^ percentile was higher in the case of Group II (20% with milk) compared to Group 1 (10% with water) (Fig 1D). However, in Groups I and II, 97% of children had higher height gain velocity than the Control, indicating that both the interventions, in water and milk, had provided gains to all the children. As seen before, gender did not influence the distribution in height gain velocity (S2B-S2C Figs). The Mann–Whitney U test for height velocities of Group I & II compared to Control have shown statistical significance (p-value<0.05) for all observed outcomes.

Gain velocities were also calculated for BMI, fat mass, fat-free mass and BMC. A 1.25 and 2 fold higher BMI gain velocities were observed for Group I and II, respectively, compared to the Control group (0.04 ± 0.06 kg/(m^2^ x month)), exhibiting a marginal overall increase in the BMI in the population. This can be rationalised as height, and weight significantly increased in the population. As expected, the BMI did not change significantly in the groups (S3A-S3B Figs). While the BMI did not alter significantly, Fig 2A shows that the fat mass gain velocities had 1.77- and 2.75-fold higher mean values for Group I and II, respectively, than the Control group (S4A-S4C Figs).

**Fig 2:**
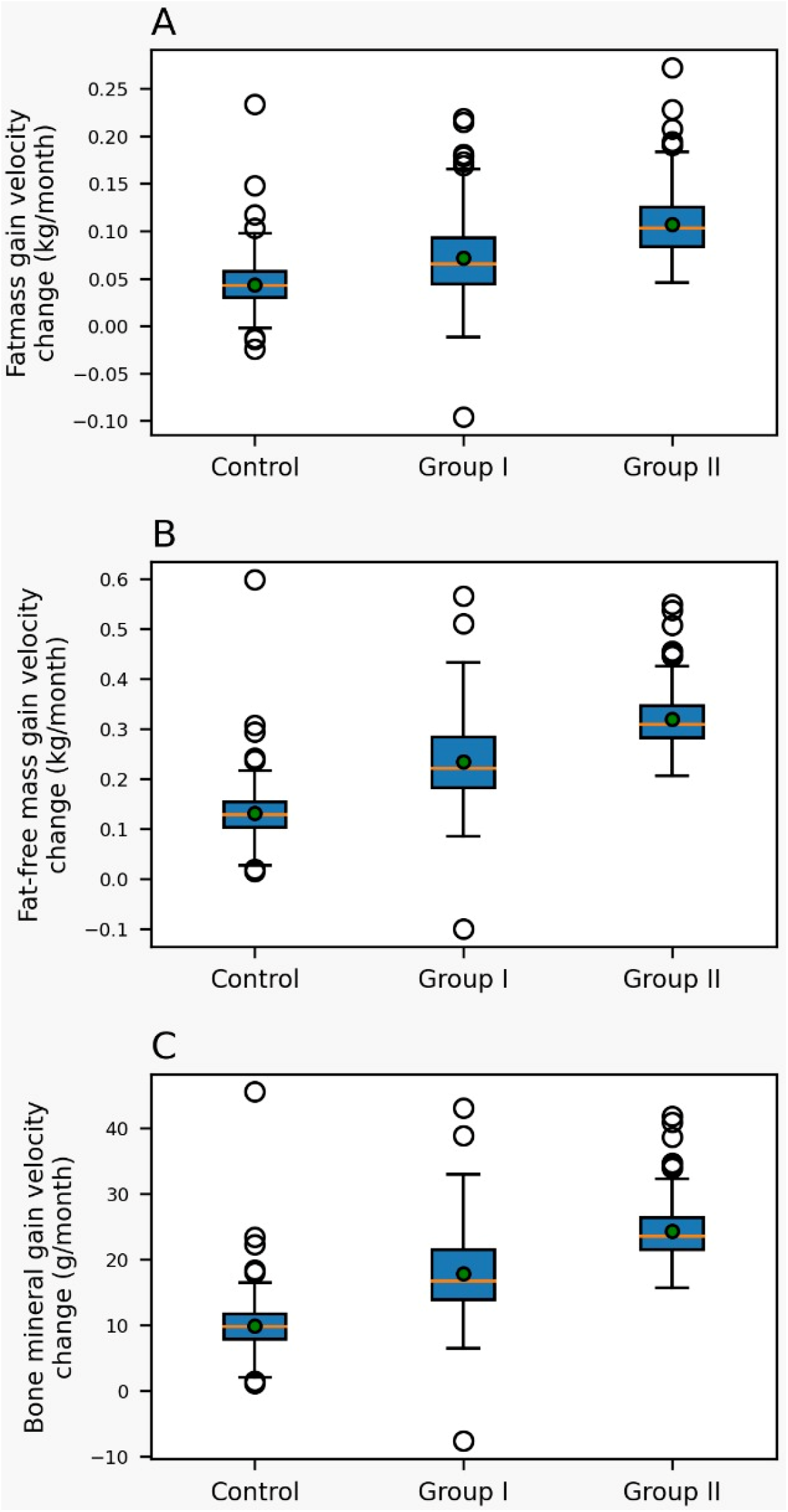
Fat mass, fat-free mass, and bone mineral content (BMC) gain velocities in Control, Group I and II children. (A) Fat mass gain velocity, the mean fat mass gain velocity of Group I, II and Control were (0.07 ± 0.04), (0.11 ± 0.03) and (0.04 ± 0.02) kg/month, respectively. (B) Fat-free mass gain velocity, the mean fat-free mass gain velocity of Group I, II and Control were (0.23 ± 0.08), (0.32 ± 0.05) and (0.13 ± 0.05) kg/month, respectively. (C) Bone Mineral Content (BMC) gain velocity, the mean BMC gain velocity for Group I, II and Control were (17.88 ± 5.90) and (24.36 ± 4.05) and (9.94 ± 3.73) g/month. Green dots indicate the mean values.

Further, Fig 2B represented changes in fat-free mass gain velocities, where 80% and 95% of children had higher fat-free mass gain velocities for Group I and II, respectively, compared to the Control group (0.13 kg/month) (S5A-S5C Figs). A significant improvement was also observed in the BMC gain velocities (Fig 2C) with 1.83- and 2.67-fold higher mean values for Group I and II, respectively, as compared to the Control group ((17.88 ± 5.90 g/month) (S6B-S6C Figs). No gender biases were observed for any gain velocities as the interventions had a comparable effect on the overall population. The Mann–Whitney U test for fat mass, fat-free mass, and BMC velocities of Group I & II compared to Control have shown statistical significance (p-value<0.05) for all observed outcomes.

The above improvements in height, weight and body composition were independently evaluated through MUAC and physical performance. Fig 3A shows 3.33- and 4.0-fold higher MUAC gain velocities in Group I and II, respectively, compared to the Control group (0.03 ± 0.11 cm/month), clearly showing a significant intervention effect on the population (S7A-S7B Figs). Both the groups showed about a 3.5-fold increase in mean MUAC values. While analysing the physical performance time, 73% and 83% of the children had significantly improved performance mean time taken by Group I and II, respectively, compared to the Control group (0.03 seconds) (Fig 3B and S8A-S8C Figs). Further, Group I showed a drop of 0.12 ± 0.09 seconds in performance time, whereas Group II showed a drop of 0.16 ± 0.1, compared to an increase of 0.03 ± 0.09 seconds in the Control group. These results clearly indicated that the overall gain in the anthropometric values was reflected in the improvements in physical performance and MUAC values.

**Fig 3:**
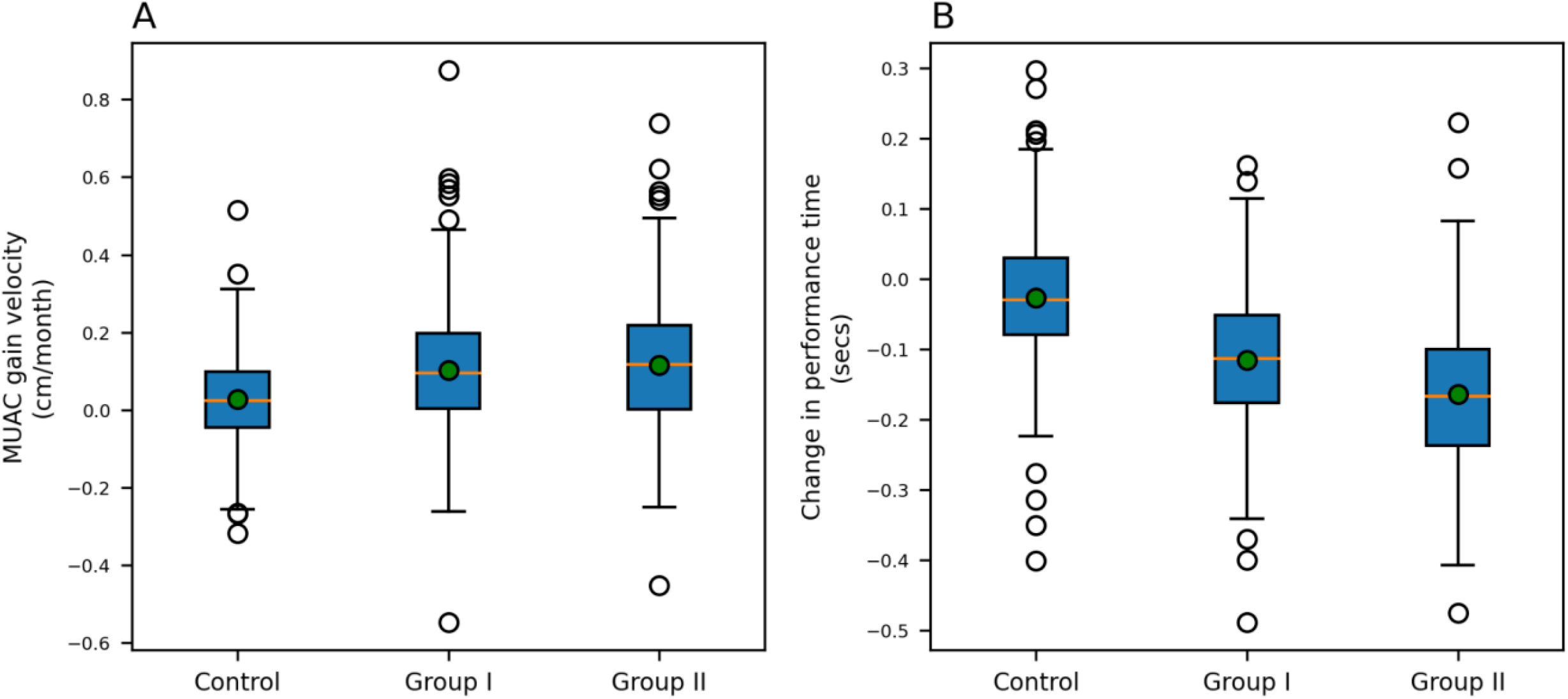
MUAC gain velocity and physical activity performance time in Control, Group I and II. (A) The MUAC gain velocity and (B) The mean change in time taken to complete the 50m dash for Control, Group I & II, the green dot indicates the mean value. The MUAC gain velocities for Group I, II and Control were (0.1 ± 0.17), (0.12 ± 0.17), and (0.03 ± 0.11) cm/month respectively, while the meantime change for Group I, II and Control were (−0.12 ± 0.09), (−0.16 ± 0.1) and (−0.03 ± 0.09) seconds respectively.

### Clinical analysis

The micronutrients present in the interventional drink would also improve children’s overall health, which was estimated by evaluating the existence of anaemia and morbidity in the children population. Fig 4A shows a significant decrease in children under the moderate and severe anaemia categories (15.7% and 5.7%) for Group I and (21.3% and 2.3%) for Group II, as compared to the Control group (6.7% and 2%). This resulted in about 22% of the children moving to the normal and mild category from moderate and severe in Group I and Group II, while only about 9% shifted to a healthier state in the Control. Fig 4B exhibits the overall changes in children’s morbidity degree scores due to the interventions. There was a significant decrease in the second and third-degree morbidity categories (34% and 11%) for Group I and (33% and 5.9%) for Group II, compared to the Control group (3.2% and 6.7%). Also, more children moved towards the morbidity-free and first-degree morbidity categories (26.1% and 18.9%) in Group I and (36.3% and 2.6%) in Group II, exhibiting a significant improvement compared to the Control group (3.9% and 7.1%), suggesting the advantage of the intervention in health status.

**Fig 4:**
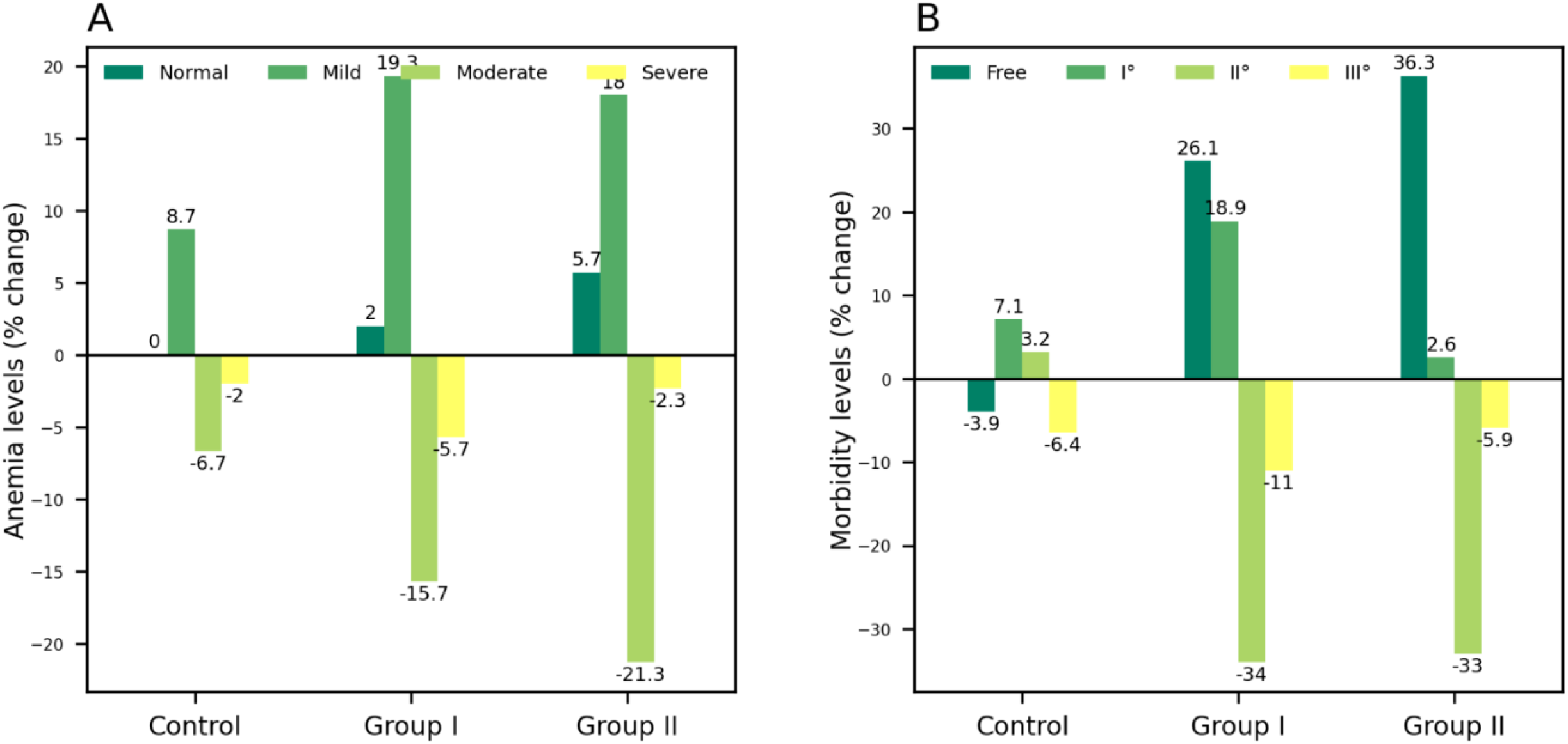
Percentage change in children with anaemia and morbidity levels in Control, Group I and II. Percentage change in children with (A) anaemia and (B) morbidity in Control, Group I and Group II. The darker green to yellow colour shows the increasing order of severity in anaemia and increasing level of morbidity starting from normal.

### Cognitive development analysis

The cognitive analysis consisted of different tests such as the CPM test, Arithmetic Test, Digit Span, and Digital Symbol test to assess various measures of cognition such as levels of intelligence, numerical ability, memory, and concentration, respectively. The mean relative score changes in the CPM test exhibited a significant change of 0.19 and 0.22 in Group I and II, respectively, compared to the Control, which was 0.09 (Fig 5A). The Mann–Whitney U test for relative score changes in the CPM test of Group I & II compared to Control have shown statistical significance (p-value<0.05). In Group I, the percentage of children with below-average, average, above-average grades decreased by 5.2%, 8.3%, and 2.4%, respectively, and shifted to high (10.1%) and very high (5.9%) grade categories. In Group II, a 4.8% and 17.8% decrease was observed in below average and average grade children, respectively, whereas the percentage of the above average, high, and very high-grade children increased by 5.2%, 12.2%, and 5.9%, respectively (Fig 5B). A 2.11 and 2.44 relative score fold change is observed in Group I and II, respectively, compared to the Control group, indicating improvement in the children’s intelligence level.

**Fig 5:**
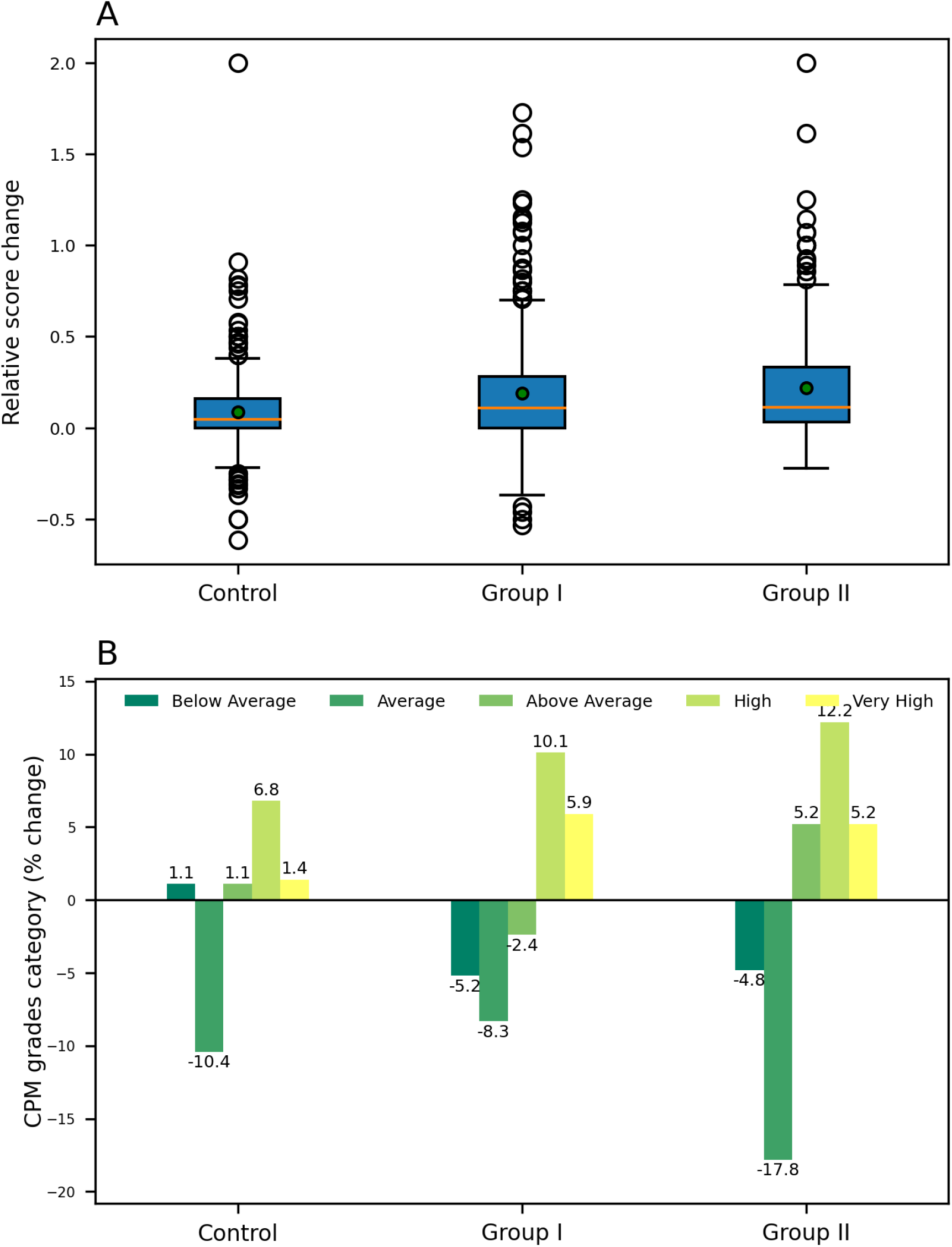
CPM test relative score change and percentage change in children’s test grades distribution in Control, Group I and II. (A) Changes in CPM test relative score, the mean relative score changes in Group I and II were 0.19 and 0.22, respectively, compared to Control (0.09), green dots indicate the mean values. (B) Percentage change in children’s CPM test grade distribution in Control, Group I and II.

Similarly, the mean relative score changes in the Arithmetic test were 0.11 and 0.15 in Group I and II, respectively, compared to the Control, which was found to be 0.08 (Fig 6A). The Mann– Whitney U test for relative score changes in the Arithmetic test of Group I & II compared to Control have shown statistical significance (p-value<0.05). The percentage of children with average grades decreased by 20.3% and 25.6% for Group I and II respectively, and change in above average and high-grade categories was observed with an increase of (19.2% and 2.1%) for Group I and (21.1% and 5.9%) for Group II, respectively (Fig 6B). In the Control group, the children with above-average grades increased by only 10.1% and showed no increase in the number of children with high grades. A 1.38 and 1.88 relative score fold change is observed in Group I and II, respectively, compared to the Control group.

**Fig 6:**
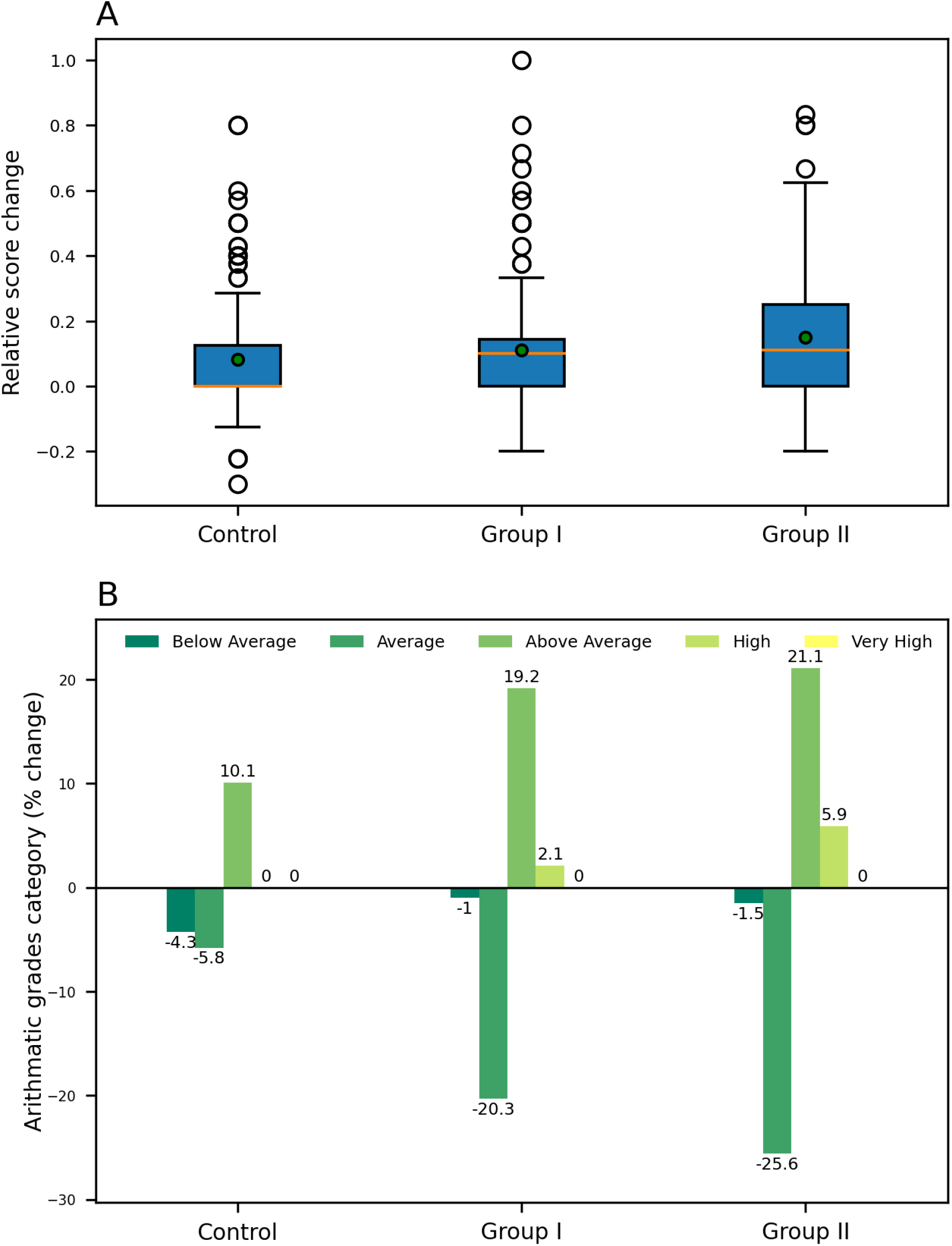
Arithmetic test relative score change and percentage change in children’s test grades distribution in Control, Group I and II. (A) Changes in the Arithmetic test relative score, the mean relative score changes in Group I and II were 0.11 and 0.15, respectively, compared to Control (0.08), green dots indicate the mean values. (B) Percentage change in children’s arithmetic test grade distribution in Control, Group I and II.

The mean relative score changes in the Digit span test of the Control group were 0.09, whereas the score changes for Group I and II are 0.14 and 0.18, respectively (Fig 7A). The Mann– Whitney U test for relative score changes in the Digit span test of Group I & II compared to Control have shown statistical significance (p-value<0.05). In the Control group, the children with above-average grades increased only by 21.2% and no increase was seen in the number of children with high grades. Whereas an increase of (19.5% and 2.8%) for Group I and (34.4% and 5.6%) for Group II was observed in above average and high-grade categories, respectively. A decrease was observed in below-average grade children across all three study groups (Fig 7B). A 1.56 and 2.0 relative score fold change was observed in Group I and II, respectively, compared to the Control group, indicating benefits of the intervention on the population.

**Fig 7:**
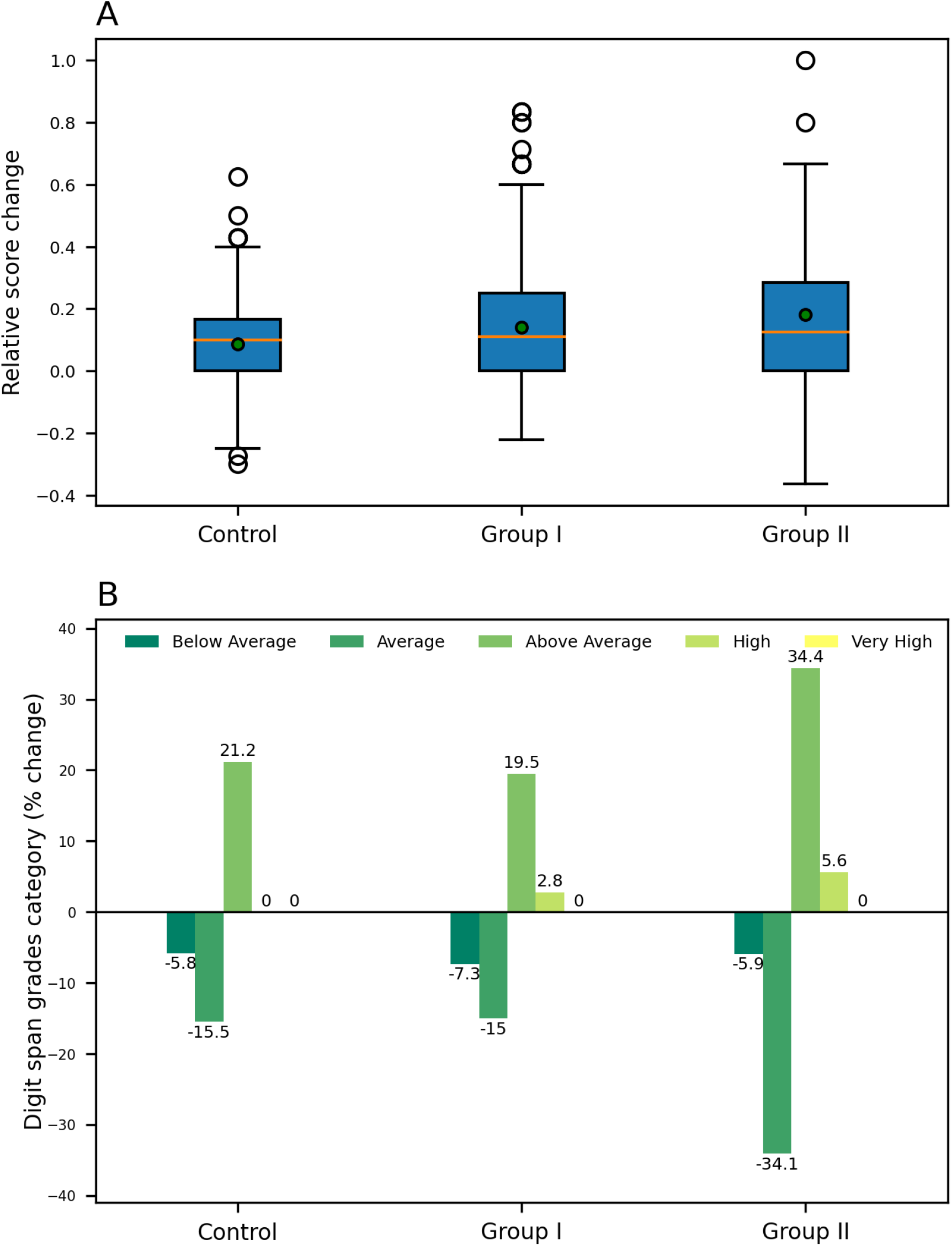
Digit Span test relative score change and the percentage change in test grades distribution in Control, Group I and II. (A) Changes in the Digit Span test relative score, the mean relative score changes in Group I and II were 0.14 and 0.18, respectively, compared to Control (0.09), green dots indicate the mean values. (B) Percentage change in children’s Digit Span test grade distribution in Control, Group I and II.

The Digit symbol test relative mean score changes of the Control group were 0.25, whereas the score changes for Group I and II were 0.31 and 0.32, respectively (Fig 8A). Although the outcome of Mann–Whitney U test has shown insignificance (p-value>0.05), the results of the grade analysis (Fig 8B) indicate that children’s concentration has been improved. Children with above-average grades increased by only 15.0% for the Control group, whereas 21.0% and 6.5% increase was observed for Group I in above-average and high-grade categories, respectively. The percentage of children with below-average and average grades decreased by (15.1% and 13.4%) for Group I and by (12.7% and 19.0%) for Group II. In Group II, the percentage of the above average, high, and very high grade’s children increased by 22.4%, 7.5%, and 1.9%, respectively. A greater decrease is observed in below-average grade’s children across all three study groups (Fig 8B). A 1.24 and 1.28 relative score fold change was observed in Group I and II, respectively, compared to the Control group, indicating improvement in children’s concentration.

**Fig 8:**
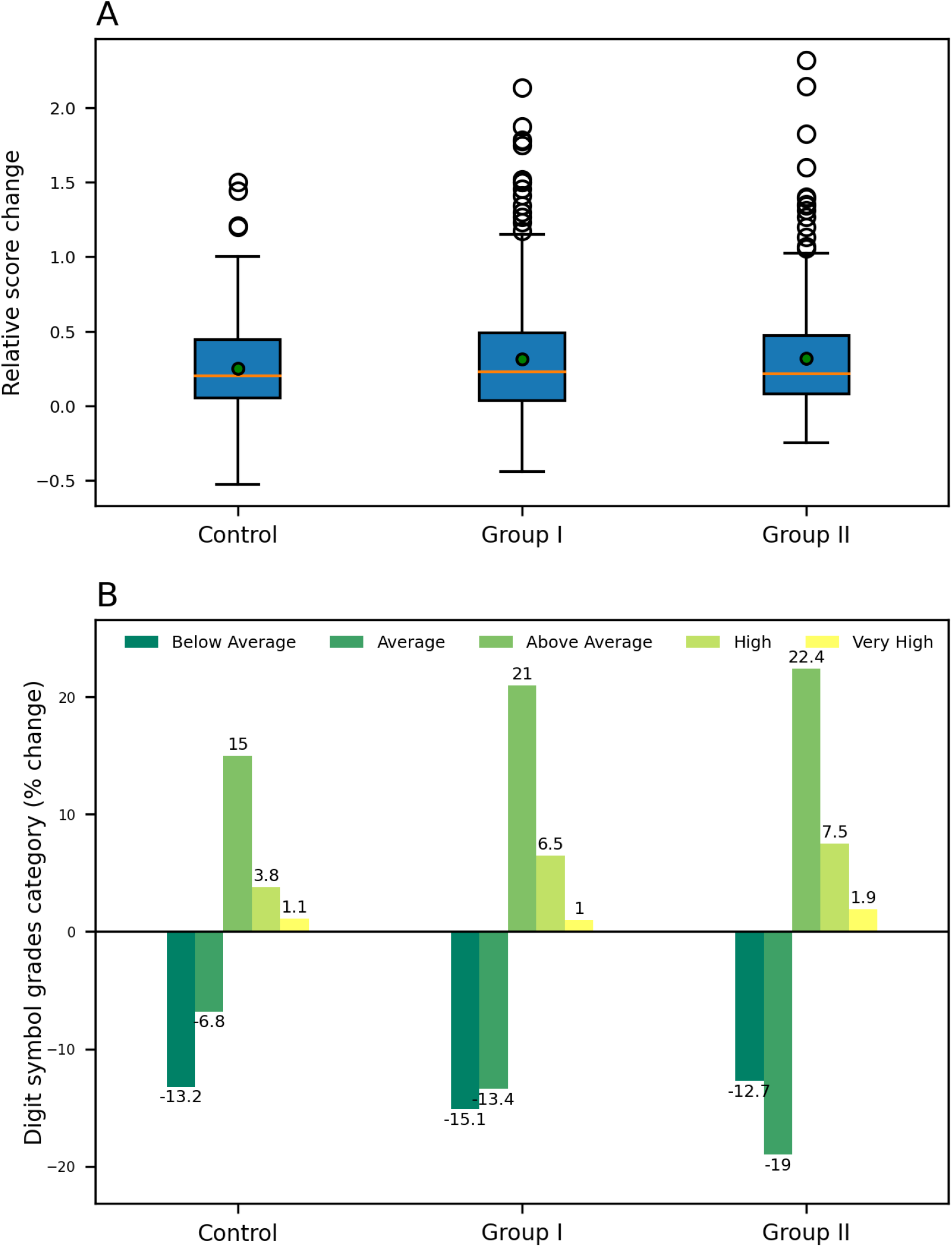
Digit Symbol test relative score change and the percentage change in test grades distribution in Control, Group I and II. (A) Changes in the Digit Symbol test relative score, the mean relative score changes in Group I and II were 0.31 and 0.32, respectively, compared to the Control (0.25), green dots indicate the mean values. (B) Percentage change in children’s Digit Symbol test grade distribution in Control, Group I and II.

## Discussion

Growth in children seems continuous when observed across years with periods of spurts and stasis. Both macronutrients and micronutrients are essential for children’s balanced nutrition. Various studies have demonstrated the importance of different nutrients on physical growth, cognitive development, and their role in physiology involving growth hormone signalling during the growth and development process [24]. Multiple nutrient interventions have received much attention as a cost-effective and promising strategy to improve child health. Supplementation or intervention with five or more nutrients has shown much stronger benefits than a single nutrient. Even more importantly, the timing of supplementation, number of nutrients, and some specific types of nutrients are associated with better effectiveness. Various nutrient or multi-micronutrient intervention studies conducted during children’s growing years have shown highly significant, positive responses on different anthropometric parameters, including linear growth and body height and weight increments in the infants and growing adolescent years [25-30]. An Indian study with over 100 girls in the age range of 10-16 years reported a weight gain velocity of 0.49kg/month as an effect of supplementation by 100 gms of biscuits [31]. The biscuits were fortified with vitamin A, iron, folic acid, vitamin C, and iodine and provided 497 kcal and 11.36 gms of protein (five days a week). The results observed are similar to our intervention, where the children exhibited a weight gain velocity of 0.43 kg/month, where the intervention provided 420 kCal and comparable protein and micronutrient content.

The micronutrients are extensively utilised for tissue growth leading to overall physical development, including weight and height [32]. A multimicronutrient study by Sazawal et al. in 2013 reported a height gain velocity of 0.455 cm/month due to a 1-year intervention (fortified yoghurt) [33]. In the present analysis, we have observed the height gain velocities to be 0.54 and 0.66 cm/month. Comparing the composition of the intervention’s two studies indicate that the milk-based micronutrient-fortified drink has 2.7x Iron, over 6x Calcium, 1.8x Iodine, 1.5x Vitamin A, and over 5x protein concentration. Another research focused on 7-9 yrs old children reported a height gain velocity of 0.6 cm/month, which is almost similar to the change reported in the present study [34]. A composition comparison indicates equivalent calorie and protein intake and critical micronutrient supplementation.

Several studies have reported associations between the multi micronutrient-enriched interventions including Calcium, Phosphorus, Vitamins A, B, C, D, and K with body composition, bone mineral content (BMC), bone area, and bone mineral density in school-aged children, which are required to maintain the integrity and mineralisation of bones. In an intervention reported by Bonjour et al., supplementation of 880 mg/d of Calcium and 380 mg/d of Phosphorus led to BMC gain velocity of 20 g/month [35]; similarly, another study with supplementation of 500mg of Calcium reported a BMC gain velocity of 15g/month [36]. In our study, we have observed the BMC gain velocity as 24g/month in Group II, which was supplemented with 888mg/d of Calcium (528g from the health drink + 360g from milk), 515mg/d of Phosphorus along with additional micronutrients, such as Vitamin D (2.4 mcg/d), Magnesium (46.2 mg/d), Vitamin K (29.70 mg/d), Vitamin C (19.8 mg/d), Copper (0.35 mg/d), Zinc (2.9 mg/d) and Manganese (0.99 mg/d), which are known to contribute to bone health [37]. These results clearly indicate the beneficial role of multi-micronutrient supplementations on a child’s BMC.

The research evidence from intervention studies suggests that high zinc concentration in the brain relates to cognitive functioning. Several B-complex vitamins are needed to synthesise several neurotransmitters. Iodine is essential for synthesising triiodothyronine and thyroxine [38, 39]. Iron is necessary for the functioning of the neurotransmission system by producing dopamine, serotonin, and GABA. Therefore, the possible mechanisms of the major nutrients can be associated physiologically as well. A recent study on 8-14 year old children indicated a 41% improvement in cognitive abilities due to micronutrient fortified drinks [40]. The current study has shown an 87% improvement in Group II, where the calorie intake is similar to the published study. A deep dive in the micronutrient content indicates a higher micronutrient content, including similar zinc concentrations, 1.6x Iron, 6x Magnesium, 25x Potassium, 3x Calcium, and Iodine (absent in the [40] study).

Research has proved that micronutrient-rich interventions have important benefits for child survival and health that justify their implementation. More comprehensive approaches that improve the diets of small children are needed to promote child growth and development [41]. Thus, all efforts should ensure that children take a well-balanced nutritious diet. Also, to identify the optimal supplementation, future studies should include both short- and long-term assessments on children’s physical, cognitive, and immune growth and development and consider the potential interactive effects of different nutrients.

## Conclusions

The study aimed to investigate a comprehensive overview and impact of the milk-based micronutrient fortified formulation on overall growth and development in Indian school-aged children. The focus was to associate various growth parameters with holistic insight, including physical performance, cognitive development, anthropometric parameters, morbidity, and different clinical outcomes. The growth process in a healthy child is accompanied by an increase in body size or mass and cognitive development at varying rates; therefore, the rate-based analysis method was used to analyse different parameters in extended time and was also compared with the specific IAP standard growth charts to be precise. It is observed that the formulation helped children in physical and cognitive development with improved clinical outcomes. Gender-specific analysis revealed no significant gender biases of the effect of formulation on different growth parameters. The results showed a significant positive effect on all the analysed growth parameters in both the intervention groups. This study demonstrates that 2 servings of health dink daily bridged the nutrient gap, thus helping children improve their nutritional status and achieve growth potential.

## Data Availability

All data produced in the present work are contained in the manuscript.

## Declarations

### Ethics approval and consent to participate

Zydus Wellness R&D team’s approval was received to use the data in the present. The dataset of the previously conducted studies was provided by Zydus Wellness R&D team, who own the dataset, on which the present analysis has been performed and the authors of current study does not require other administrative permission except Zydus Wellness R&D team. The data provided to us by Zydus Wellness team was in the anonymized format.

The original study was performed as per accordance with the Declaration of Helsinki. All relevant guidelines and regulations under ethics approval and consent to participate section in the declaration were followed while conducting the study. An approval from the Avinashilingam University’s ethics Committee was obtained for the study. Meetings were conducted with parents of all participating children to inform them about the study and obtained written consent. Ethics committee meeting was held on 5th July 2006 and an approval no. HEC.2006.01 was provided for the study.

Present study, is the secondary analysis of the original study published as below:

1. Vijayalakshmi P, Premakumari S, Haripriya S. Supplementation of Milk Based Health Drink Enriched with Micro Nutrients-Part-I Impact on Growth and Haemoglobin Status of 7-12 Year Old Children. The Indian Journal of Nutrition and Dietetics. 2008 Nov 1;45(11):449-65.
2. Vijayalakshmi P, Premakumari S, Haripriya S. Supplementation of Milk Based Health Drink Enriched with Micronutrients Part II - Impact on Clinical and Morbidity Picture, Physical Performance and Cognitive Development of 7-12 Year Old Children [Internet]. Informaticsjournals.com. 2021 [cited 11 October 2021]. Available from: http://www.informaticsjournals.com/index.php/ijnd/article/view/4896

### Consent for publication

Not applicable

### Availability of data and materials

All data generated or analyzed during this study are included in this article and the Additional file.

### Authors’ contributions

KL analyzed and interpreted the data. VG and SR were a major contributor in writing the manuscript. KVV was responsible for guidance and validation. All authors read and approved the final manuscript.

## Acknowledgements

The authors acknowledge the support from the Zydus Wellness R&D team for providing resources including the study dataset.

