## Additional file for "Effect of a 12-mo milk-based micronutrient-fortified drink intervention on children: a systemic analysis of placebo-controlled study dataset"

[*]MetFlux Research Private Limited

[+]Professor, Dept. of Chemical Engineering, IIT Bombay

### **Materials and Methods.** Extended version.

#### Intervention

S1 Table provides the nutritional composition of 66 grams of micronutrient-fortified formulation and 300 ml of toned milk. In the case of Group I, the 33 grams of the formulation was given in 150 ml of water twice a day, while Group II was given 33 grams of micronutrient-fortified drink in 150 ml of toned milk twice every day.

**S1 Table: The nutritional composition of 66 grams of micronutrient-fortified formulation and 300 ml of toned milk.**

| Nutrients | Formulation | Toned Milk | Nutrients | Formulation | Toned Milk |
|---|---|---|---|---|---|
| Energy (kcal) | 276 | 201 | Vitamin B12 (mcg) | 0.50 | 0.047 |
| Milk protein (g) | 11.88 | 9.0 | Folic acid (mcg) | 50 | - |
| Fat (g) | 7.26 | 10.2 | Vitamin A (I.U) | 700 | 17.7 |
| Carbohydrate (g) | 41 | 15.0 | Vitamin E (mg) | 1.98 | - |
| Calcium (mg) | 528 | 360 | Vitamin C (mg) | 19.8 | 6.0 |
| Phosphorus (mg) | 514.8 | 270 | Zinc (mg) | 2.97 | - |
| Vitamin D (I.U.) | 99 | - | Vitamin B1 (mg) | 0.30 | 0.12 |
| Sodium (mg) | 264 | 57 | Vitamin B2 (mg) | 0.38 | 0.3 |
| Potassium (mg) | 607 | 270 | Vitamin B6 (mg) | 0.50 | 0.03 |
| Chloride (mg) | 330 | - | Niacinamide (mg) | 3.96 | 0.3 |
| Iodine (mcg) | 73 | - | Calcium Pantothenate (mg) | 1.98 | - |
| Iron (mg) | 8.91 | 0.6 | Vitamin K (mg) | 0.03 | - |

### Assessment

#### Physical growth analysis

The anthropometric measurements such as weight, height, and MUAC are used for the physical growth analysis. Height and weight measurement data is given each month over the 12-month study period. The MUAC analysis is useful to assess undernutrition. It is a good predictor of mortality in children better than any other anthropometric indicator. The MUAC data is given every three months. The dataset includes body mass index (BMI) and physical performance time. The physical performance time is taken in seconds to cover the 50m dash [21]. The body composition parameters such as body fat mass, fat-free mass, and bone mineral content (BMC) are calculated using correlations. The bone mineral content is calculated using the reference from the literature [27]. The IAP growth percentiles are calculated for weight and height for baseline and final measurement [23]. The growth parameter velocities of all the participating children are calculated by subtracting the initial value from the final and dividing the result by 12 months (duration of the study).

#### Clinical analysis

The anaemia and morbidity level data is given for clinical outcome analysis. The baseline and final measurements are given. The degree of anaemia is calculated using blood haemoglobin levels measured through the cyan-methaemoglobin method. The blood haemoglobin level was accessed in 10% of study participants (n=90). The morbidity level data are provided in the dataset and derived using a morbidity assessment schedule [21]. The morbidity severity is calculated using a morbidity scoring pattern in a year. The anaemia severity and morbidity scoring are given in S2 and S3 Tables, respectively, under the supplementary section.

#### Cognitive development analysis

Cognitive functions comprise multiple cognitive domains such as memory, language, perception, attention, and executive functions. Several tests assess the different domains of cognitive functions in children. For example, Matrix Reasoning Test (measures nonverbal analytical reasoning based on the ability to complete patterns and analogies), Similarity Test (measures verbal rationale based on the ability to form verbal concepts), School Readiness Test (assesses the acquisition of pre-literacy and pre numeracy skills in preparation for primary school), Digit Span Test (measures attention, short term memory, and auditory sequencing), Vocabulary test (measures verbal memory), Benton visual retention test (measures the visual-spatial perception, visual and verbal conceptualization, and immediate memory span), and Cattell's retentivity test (measures the visual recall for irregular geometrical designs and delayed memory span). The cognitive assessment dataset is given for all Group I & II and control groups. The dataset comprises scores and grades for Raven's Coloured Progressive Matrices (RCPM) test for intelligence, Digit Span test for memory, Arithmetic test for numerical reasoning and concentration, and Digit Symbol for coordination and concentration. These cognitive tests were administered to all the children at baseline and the end of the supplementation period. The raw score, the total number of questions answered on a particular subtest, is converted to a percentage score and grades [21].

**Physical growth parameters**

The measured parameters in the children dataset are used to calculate growth indicators such as Body Mass Index (BMI), body fat mass, and fat-free mass are calculated using correlations given below.

$$\text{Body Mass Index (BMI)} = \text{Weight} / \text{Height}^2$$

$$\text{Body fat mass (Boys)} = (-0.05 * \text{Age}) + (1.034 * \text{BMI}) + 4.39 - (3.6 * \text{Age} / 18)$$

$$\text{Body fat mass (Girls)} = (-0.05 * \text{Age}) + (1.034 * \text{BMI}) + 4.39$$

$$\text{Fat-free mass} = \text{Body weight} - \text{Body fat mass}$$

The Bone Mineral Content (BMC) is also calculated using regression correlation [27].

**Morbidity Score**

The morbidity severity is calculated using a morbidity scoring pattern in a year. The anaemia severity levels are calculated based on haemoglobin levels in the blood given in the S2 Table. The scoring scheme is given in the S3 Table.

**S2 Table: Degree of Anaemia**

| Hemoglobin (g/dl) | Anemia level |
|---|---|
| > 12 g/dl | Normal |
| 10-12 g/dl | Mild Anemia |
| 7-10 g/dl | Moderate Anemia |
| < 7g/dl | Severe Anemia |

**S3 Table: Morbidity Scoring Pattern**

| Morbidity Details | Scores given |
|---|---|
| **Diarrhoea** | |
| 15 days or more | 30 |
| 7-14 days | 20 |
| 4-6 days | 10 |
| 1-3 days | 5 |
| **Fever** | |
| 10 days or more | 20 |
| 5-9 days | 10 |
| 1-4 days | 5 |
| **Cold** | 5 |
| **Other morbidities** | |
| For 10 days or more | 20 |
| Within 10 days | 10 |

### List of figures

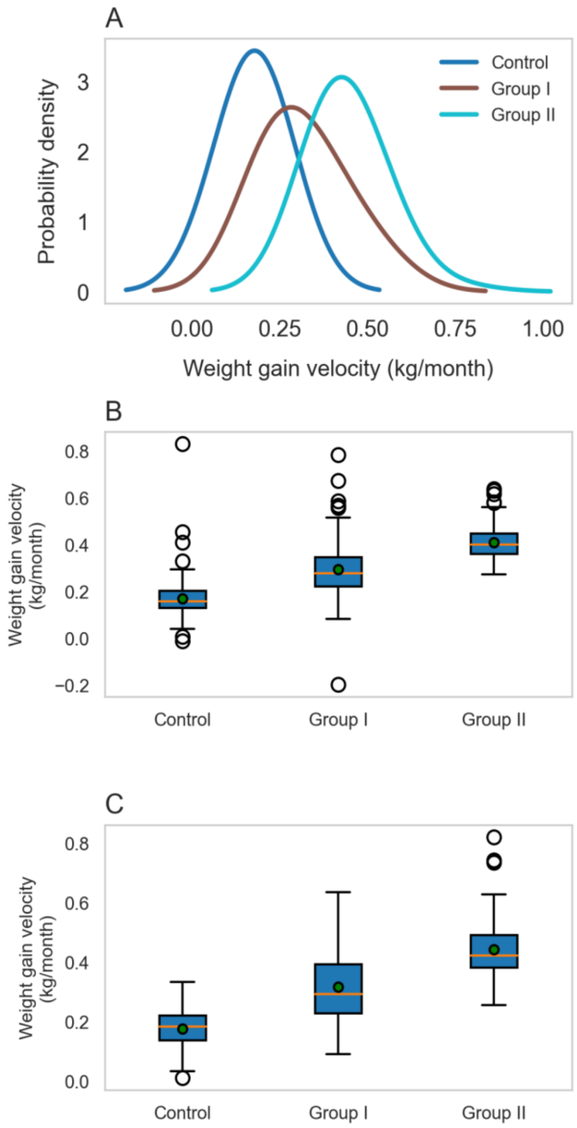


**S1 Fig. Probability distribution of weight gain velocities and gender-specific effect on Control, Group I and II children.** (A) The probability distribution of weight gain velocities is shown for Control, Group I, and II. Group I and II curves are shifted toward the right, showing higher weight gain velocities than the Control group. (B) The weight gain velocity is shown for boys in Control, Group I & II, where the green dot indicates the mean value. The weight gain velocities for the Group I and II are (0.29 ± 0.11) and (0.41 ± 0.07), respectively, which are considerably higher than the Control group (0.17 ± 0.08) kg/month. (C) The weight gain velocities for the girls the Group I and II are (0.31 ± 0.11) and (0.44 ± 0.09) respectively, which are considerably higher than the Control group (0.17 ± 0.05) kg/month.

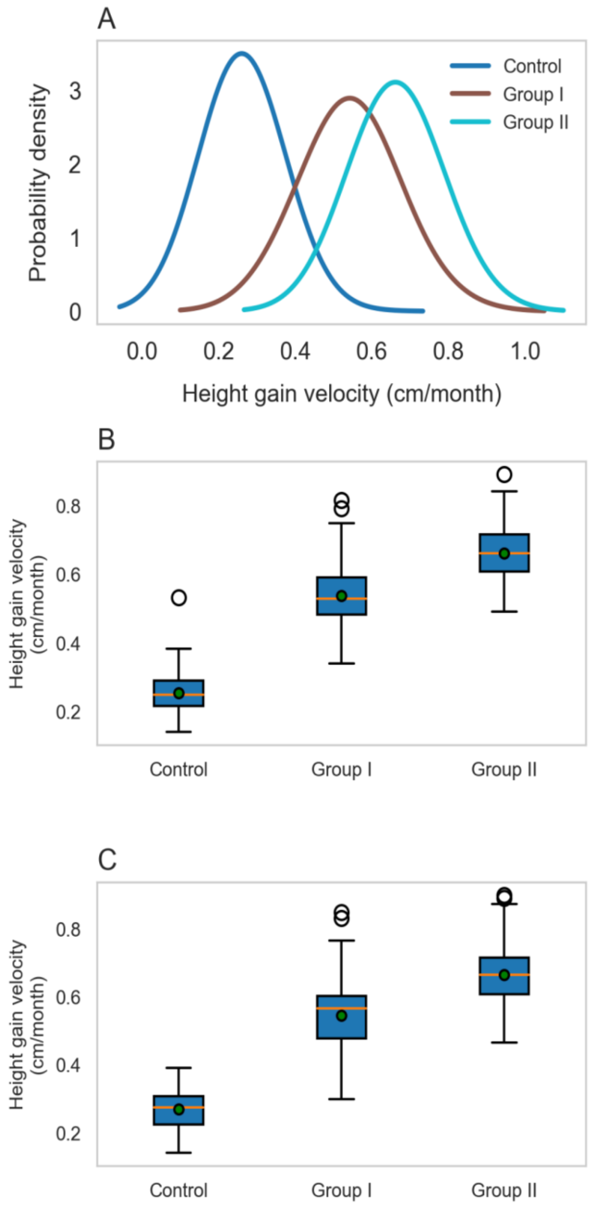


**S2 Fig. Probability distribution of height gain velocities and gender-specific effect on Control, Group I and II children.** (A) The probability distribution of height gain velocities is shown for Control, Group I, and II. Group I and II curves are shifted toward the right, showing higher Height gain velocities than the Control group. (B) The height gain velocity is shown for boys in Control, Group I & II, where the green dot indicates the mean value. The height gain velocities for the Group I and II are (0.54 ± 0.10) and (0.66 ± 0.08), respectively, which are considerably higher than the Control group (0.27 ± 0.05) cm/month. (C) The height gain velocities for the girls in Group I and II are (0.55 ± 0.11) and (0.67 ± 0.09) respectively, which are considerably higher than the Control group (0.27 ± 0.05) cm/month.

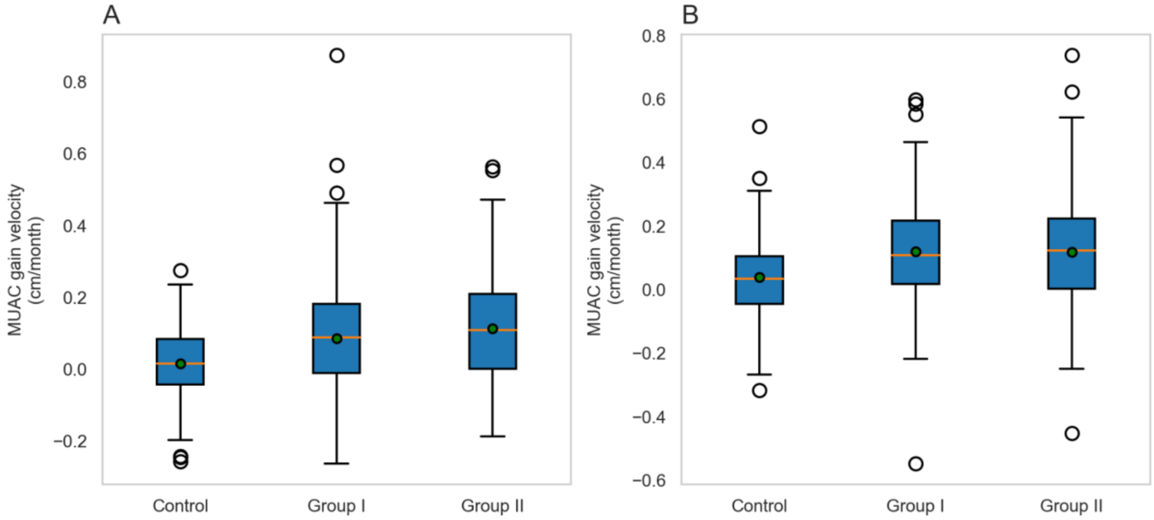


**S3 Fig. Probability distribution of Body Mass Index (BMI) gain velocities and Control, Group I and II children.** (A) The probability distribution of BMI gain velocities is shown for Control, Group I, and II. The curve of Group II is shifted toward the left, showing improved BMI compared to the Control and Group I children. (B) The BMI gain velocities are shown for Control, Group I & II, where the green dot indicates the mean value. The mean BMI gain velocities for Group II is (0.09 ± 0.05), which is considerably higher than (0.04 ± 0.04), (0.04 ± 0.06) kg/m$^2$ x month for Control and Group I, respectively.

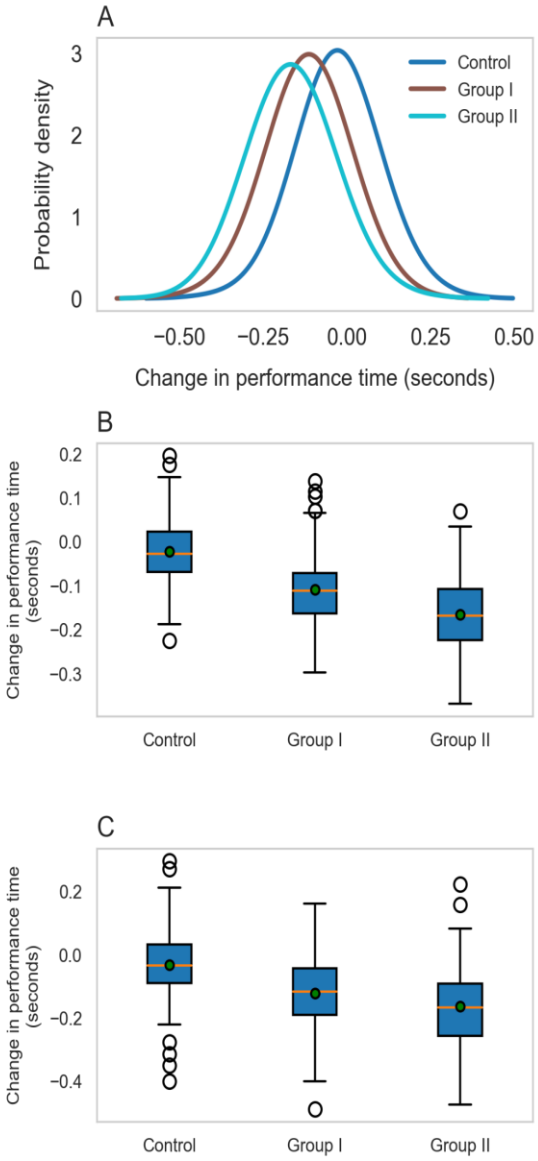


**S4 Fig. Probability distribution of body fat mass gain velocities and gender-specific effect on Control, Group I and II children.** (A) The probability distribution of body fat gain velocities is shown for Control, Group I, and II. Group I and II curves are shifted toward the right, showing higher fat mass gain velocities than the Control group. (B) The body fat gain velocity is shown for boys in Control, Group I & II, where the green dot indicates the mean value. The fat mass gain velocities for the Group I and II are (0.06 ± 0.03) and (0.1 ± 0.02), respectively, which are considerably higher than the Control group (0.04 ± 0.02) kg/month. (C) The fat mass gain velocities for girls in Group I and II are (0.08 ± 0.04) and (0.12 ± 0.03), respectively, which are considerably higher than the Control group (0.05 ± 0.02) kg/month.

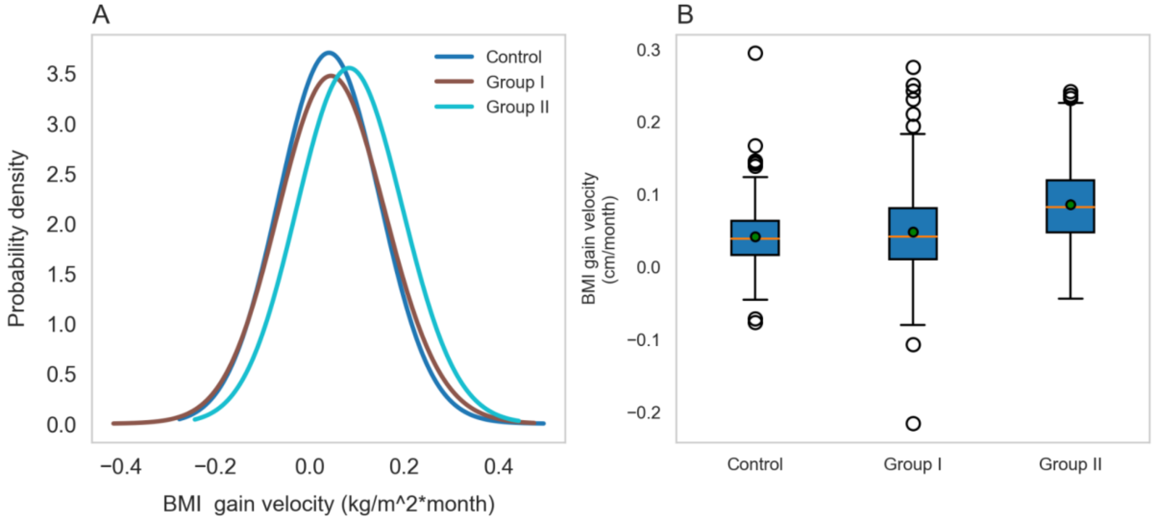


**S5 Fig. Probability distribution of fat-free mass gain velocities and gender-specific effect on Control, Group I and II children.** (A) The probability distribution of fat-free mass gain velocities is shown for Control, Group I, and II. Group I and II curves are shifted toward the right, showing higher fat-free mass gain velocities than the Control group. (B) The fat-free mass gain velocity is shown for boys in Control, Group I & II, where the green dot indicates the mean value. The fat-free mass gain velocities for the Group I and II are (0.23 ± 0.08) and (0.31 ± 0.05) respectively, which are considerably higher than the Control group (0.13 ± 0.06) kg/month. (C) The fat-free mass gain velocities for girls in Group I and II are (0.24 ± 0.07) and (0.32 ± 0.06) respectively, which are considerably higher than the Control group (0.13 ± 0.03) kg/month.

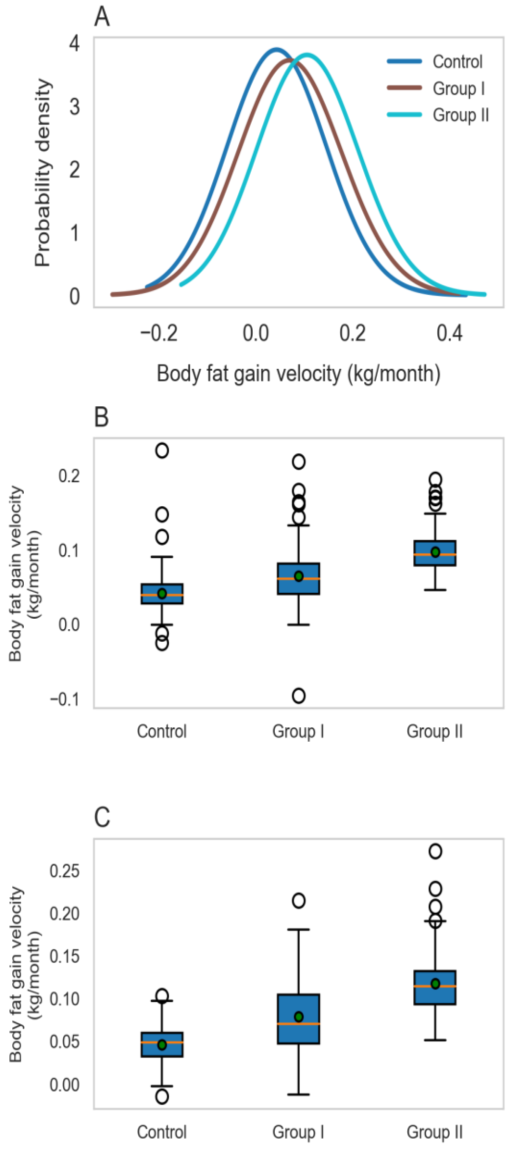


**S6 Fig. Probability distribution of Bone Mineral Content (BMC) gain velocities and gender-specific effects on Control, Group I and II children.** (A) The probability distribution of Bone Mineral Content (BMC) gain velocities is shown for Control, Group I, and II. Group I and II curves are shifted toward the right, showing higher BMC gain velocities than the Control group. (B) The BMC gain velocity is shown for boys in Control, Group I & II, where the green dot indicates the mean value. The BMC gain velocities for the Group I and II are (0.11 ± 0.04) and (0.15 ± 0.02), respectively, which are considerably higher than the Control group (0.06 ± 0.03) kg/month. (C) The BMC gain velocities for girls in Group I and II are (0.12 ± 0.04) and (0.16 ± 0.03) respectively, which are considerably higher than the Control group (0.06 ± 0.09) kg/month.

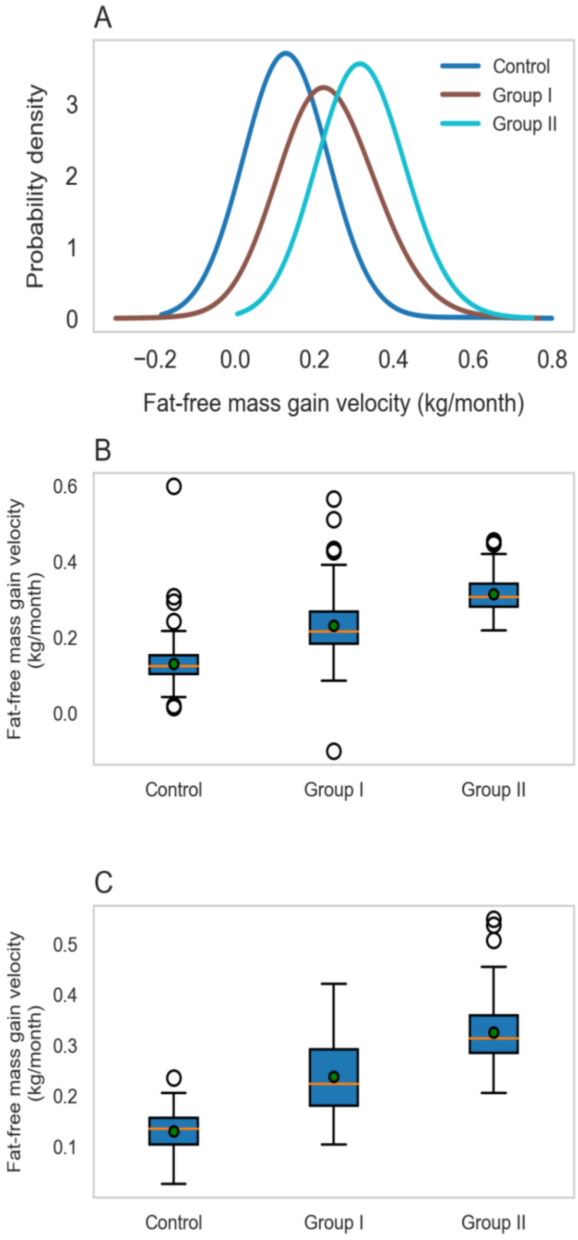


**S7 Fig. Gender-specific effect of the Mid-Upper Arm Circumference (MUAC) gain velocities on Control, Group I and II children.** (A) The MUAC gain velocity is shown for boys in Control, Group I & II, where the green dot indicates the mean value. The MUAC gain velocities for the Group I and II are (0.1 ± 0.16) and (0.11 ± 0.15), respectively, which are considerably higher than the Control group (0.01 ± 0.09) cm/month. (B) The MUAC gain velocities for the girls in Group I and II are (0.12 ± 0.17) and (0.11 ± 0.18) respectively, which are considerably higher than the Control group (0.03 ± 0.12) cm/month.

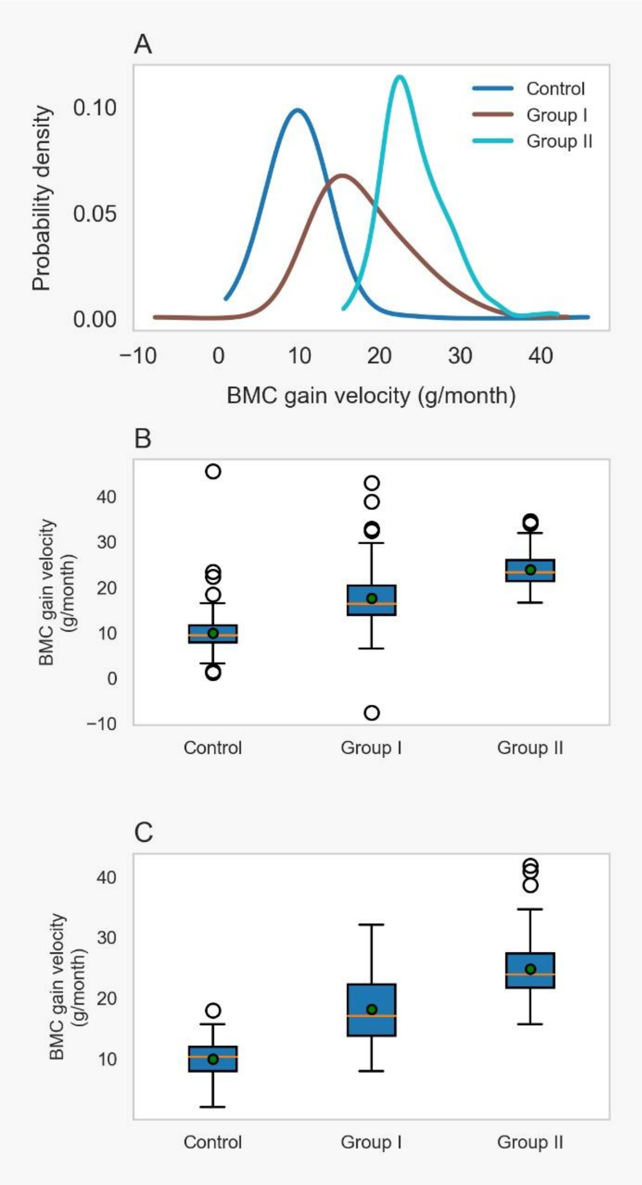


**S8 Fig. Probability distribution of change in performance time and gender-specific effect on Control, Group I and II children.** (A) The probability distribution of change in physical performance time is shown for Control, Group I, and II. Group I and II curves are shifted toward the left, showing improved physical performance compared to the Control group. (B) The change in physical performance time is shown for boys in Control, Group I & II, where the green dot indicates the mean value. The change in performance time for Group I and II are (-0.01 ± 0.07) and (-0.16 ± 0.08), respectively, which are considerably lower than the Control group (-0.02 ± 0.07) seconds. (C) The change in performance time for the girls in Group I and II are (-0.12 ± 0.10) and (-0.16 ± 0.11) respectively, which are considerably lower than the Control group (-0.03 ± 0.11) seconds.
